# Controlled evaLuation of Angiotensin Receptor Blockers for COVID-19 respIraTorY disease (CLARITY): Statistical analysis plan for a randomised controlled Bayesian adaptive sample size trial

**DOI:** 10.1101/2021.08.17.21262196

**Authors:** J. M. McGree, C. Hockham, S. Kotwal, A. Wilcox, A. Bassi, C. Pollock, L. M. Burrell, T. Snelling, V. Jha, M. Jardine, M. Jones, for the CLARITY Trial Steering Committee

## Abstract

The CLARITY trial (Controlled evaLuation of Angiotensin Receptor Blockers for COVID-19 respIraTorY Disease) investigates the effectiveness of angiotensin receptor blockers in addition to standard care compared to placebo (in Indian sites) with standard care in reducing the duration and severity of lung failure in patients with COVID-19. The CLARITY trial is a multi-centre, randomised controlled Bayesian adaptive trial with regular planned analyses where pre-specified decision rules will be assessed to determine whether the trial should be stopped due to sufficient evidence of treatment effectiveness or futility. Here we describe the statistical analysis plan for the trial, and define the pre-specified decision rules, including those that could lead to the trial being halted. The primary outcome is clinical status on a 7-point ordinal scale adapted from the WHO Clinical Progression scale assessed at Day 14. The primary analysis will follow the intention-to-treat principle. A Bayesian adaptive trial design was selected because there is considerable uncertainty about the extent of potential benefit of this treatment.

**Trial registration:** ClinicalTrials.gov, NCT04394117. Registered on 19 May 2020.

https://clinicaltrials.gov/ct2/show/NCT04394117

Clinical Trial Registry of India: CTRI/2020/07/026831

**Version and revisions:** Version 1.0. No revisions.

## 1 Introduction

The virus responsible for COVID-19, SARS-CoV-2, binds to angiotensin-converting enzyme 2 (ACE2) [14, 16]; a key regulator of the renin-angiotensin system (RAS) that is expressed on the surface of host cells. This binding may result in the downregulation of ACE2, leading to dysregulation of the RAS towards a pro-inflammatory, pro-fibrotic state [27, 24]. Another RAS protein, the angiotensin II type 1 receptor (AT1R), may play a role in COVID-19 pathophysiology by facilitating ACE2 receptor-mediated endocytosis [4] or by causing local RAS over-activation. Angiotensin Receptor Blockers (ARBs) are widely available medications originally developed as blood pressure-lowering agents that may help prevent cardiac events and the progression of kidney disease. By preventing angiotensin II from binding to AT1R, it is hypothesised that ARBs may reduce the inflammatory effects of SARS-CoV-2 infection. ARBs have been in clinical use for over 30 years and are extensively used in the treatment of chronic conditions involving a dysregulated RAS. ARBs are known to protect against lung injury in animal studies, including injury from viruses like the SARS-CoV-1 virus [15]. Whether similar protection is observed in humans is unknown.

The CLARITY trial is a two-arm, multi-centre, comparative effectiveness Phase III randomised controlled Bayesian adaptive trial, conducted in India and Australia. It is designed to evaluate whether ARBs reduce the severity of COVID-19 among high-risk patients. The investigational arm of the study is ARB delivered with standard of care. In India, the control arm is placebo plus standard care and participants are blinded. In Australia, the control arm is standard care and neither participants nor clinicians are blinded.

At the time of writing, participants are being recruited at clinical sites in India and Australia and the first planned analysis is imminent. In both countries, participants are recruited from sites providing inpatient care. In addition, in Australia, participants are recruited from sites that are managing community-based patients in monitored settings.

The trial design addresses the safety and feasibility challenges of running a clinical trial during a pandemic. To address *a priori* uncertainty about the extent of potential benefit, an adaptive sample size design has been selected that allows the trial to continue until pre-specified levels of evidence of effectiveness or futility are met. The approach reduces the risk of an indeterminate outcome and ensures the trial does not continue recruiting participants beyond the point where conclusive evidence of benefit is found. This means that results can be expeditiously reported and adopted into clinical practice. Such an approach is particularly appealing during a pandemic. In light of the well-established profile of ARBs, infection control risks, and the potential demands on health services, the burden on clinical staff is reduced through a number of measures including the collection of data largely limited to information contained in the health record. Trial specific in-person encounters are also avoided and phone consent employed to reduce opportunities for infection transmission.

The CLARITY trial statistical analysis plan (SAP) was developed following the Guidelines for the Content of Statistical Analysis Plans in Clinical Trials [8] and includes pre-specified decision rules for continuing or stopping the trial based on effectiveness or futility. None of the authors have been unblinded nor observed any data other than aggregated baseline characteristics. The final study report will follow the Consolidated Standards of Reporting Trials guidelines for reporting on randomised controlled adaptive trials [5, 22, 29]. The study protocol has been accepted for publication [13].

## 2 Study Design

### 2.1 Overview

CLARITY is a two-arm Bayesian adaptive randomised controlled trial with the sole adaptation relating to sample size. Frequent planned analyses will be performed to evaluate whether a treatment benefit exists up to a maximum sample size of 2200. Pre-specified decision rules are defined that allow the trial to be stopped at planned analyses if there is sufficient evidence of treatment effectiveness or futility.

The pre-defined decision rules for early stopping are:

1. Stop for effectiveness - if the predictive probability of trial success at the current sample size exceeds a threshold of 0.95
2. Stop for futility - if the predictive probability of trial success at the maximum sample size is below a threshold of 0.02

The first planned analysis will be triggered 14 days after enrollment of the 700th trial participant and conducted as close to that as is practical given logistical constraints. If neither the effectiveness nor futility decision rule requirements are met, successive planned analyses will be conducted 14 days after every additional 300 participants are enrolled. However, if a trial decision rule is met and the decision subsequently recommended by the Data Safety Monitoring Board (DSMB), then all enrolled participants will be followed up to 28 days after enrollment and the final analyses will be undertaken and reported on primary and secondary outcomes.

### 2.2 Intervention

In India, the ARB used is telmisartan and is supplied as 40mg tablets. In Australia, supply chain interruptions prevented the sourcing of placebo for these hygroscopic medications at the commencement of the trial. The trial was therefore commenced open-label in Australia, with the intention of sourcing placebo as the trial progressed, an intention which was subsequently abandoned with the abatement of cases in Australia and subsequent low recruitment rates. Trial Principal Investigators in Australia are permitted to select an ARB from the local hospital formulary.

The ARB or placebo is taken according to the treating clinician prescription for 28 days. Guidance is provided on the initial dose, dose titration and associated monitoring. Final decisions on ARB dosing and management are determined by treating clinicians in line with their familiarity with these medications. Participants are followed daily between Day 0 and Day 28, and at Day 90, primarily through information recorded in the medical record supplemented by phone calls.

Standard care is at the discretion of the treating team, who are encouraged to manage participants according to local best practice throughout the course of the study. Agents used for treatment of COVID-19 are determined by local practice and will be recorded.

### 2.3 Randomisation

Participants are randomised (1:1) to the investigational arm (ARB plus standard care) or control. As already noted, in India, the control arm is placebo plus standard care, and, in Australia, the control arm is standard care. Participants are randomised within 10 days after a confirmed SARS-CoV-2 diagnosis. The randomisation procedure uses permuted blocks with sizes of 4 and 6 (with equal probability). This was generated in SAS for the whole trial by Dr Qiang Li, a senior biostatistician at The George Institute for Global Health. The randomisation was stratified by country, and in Australia by hospital and community-based settings. The randomisation list and seed are held in a Redcap database and on restricted access folders that are only accessible by Qiang Li and unblinded database administrators.

### 2.4 Trial population

A flow diagram of the trial and participant progression is presented in Figure 1. Participants enrolled in the trial are those aged ≥18 years with a recent diagnosis of SARS-CoV-2 infection (*<*10 days prior to randomisation). Additional eligibility criteria identify participants who are at high risk of severe disease, including requirement for hospital admission due to COVID-19 or, for those who are managed in the community and have at least one of the following risk factors: aged ≥60 years, body mass index ≥30kg/m^2^, diagnosis of diabetes (HbA1c ≥7% and/or use of glucose-lowering medication), history of cardiovascular disease^*^, history of chronic respiratory disease^*^ or current treatment with immunosuppressive therapy (^*^as defined by the treating clinician). Additional inclusion and exclusion criteria are defined in the trial protocol [13].

**Figure 1:**
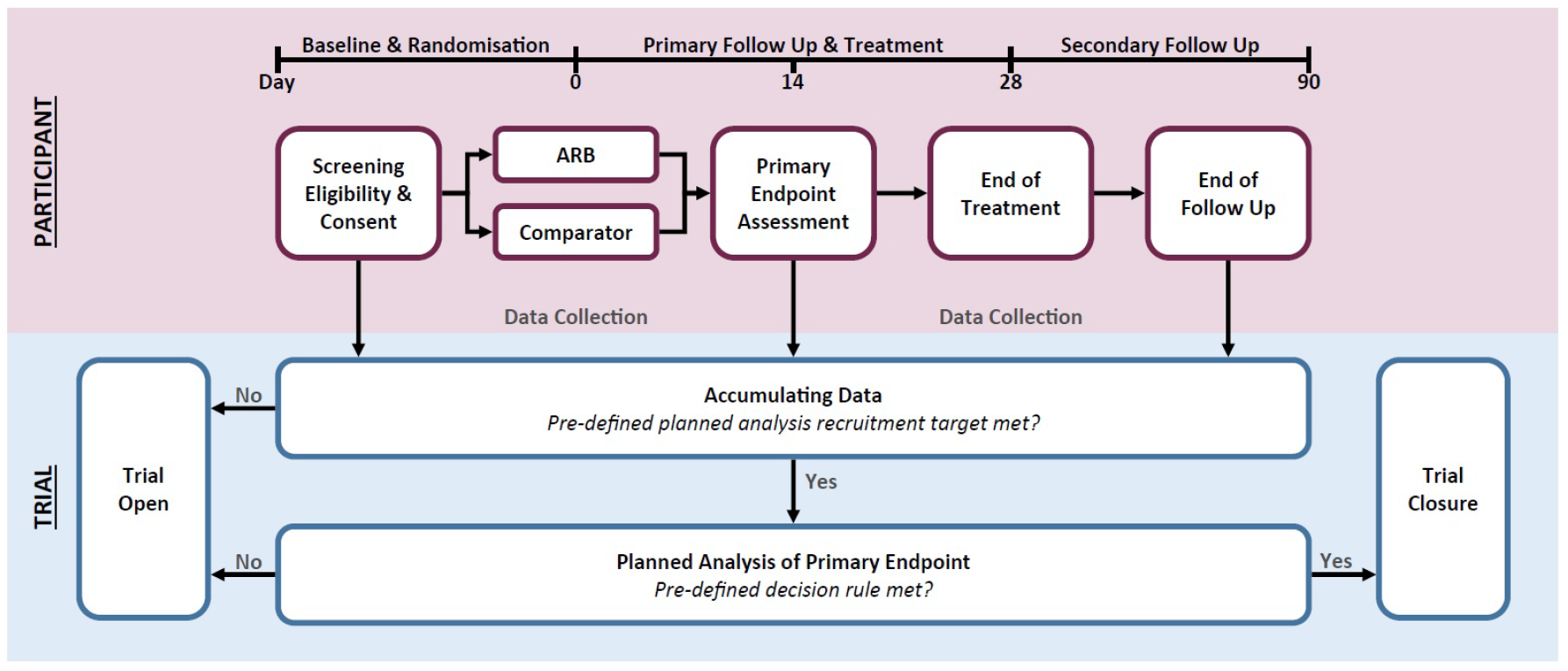
Trial overview and participant schedule [13]

The number of participants who are screened, randomised (% of screened), who complete 28 days of the trial (% of randomised) and who complete follow-up (% of 28 days completed) will be reported. The number of participants who meet the eligibility criteria but are not enrolled and the reasons for non-enrollment will be reported, where available. For missing data, it will be reported if no data have been collected from a participant for *>* 7 consecutive days between Day 0 and Day 28, the number of withdrawals (% of randomised) and those lost to follow-up (% of randomised).

### 2.5 Primary analysis population

The primary analysis population will include all participants who were randomised, and have passed the primary outcome of 14 days after randomisation with their primary outcome status either known or known to be missing. This analysis set will follow the intention-to-treat principle in that all randomised participants will be included and analysed according to the treatment to which they were initially allocated, irrespective of any deviations from this or any other protocol deviations. This analysis population will inform the primary estimand. Any missing data for the primary outcome will be reported. The occurrence of missingness will also be explored to identify any potential patterns. In the decision procedures, missing values will be handled via Bayesian imputation [28, 18].

### 2.6 Primary, secondary and exploratory outcomes

The primary outcome for this trial is a 7-point ordinal scale of clinical health status (formed by adapting the WHO Clinical Progression score [33]) assessed on Day 14. Each level in the scale is scored as shown in Table 1.

**Table 1:** 7-point ordinal scale of clinical health status [33]

| Score | Status | % Day 14 | % Day 28 |
| --- | --- | --- | --- |
| 1 | Not hospitalised, no limitations on activities | 0 | 2.3 |
| 2 | Not hospitalised, some limitation on activities | 31 | 78 |
| 3 | Hospitalised, not requiring supplemental oxygen | 34 | 1.1 |
| 4 | Hospitalised, requiring supplemental oxygen | 15.5 | 2.8 |
| 5 | Hospitalised, requiring non-invasive mechanical ventilation or high flow nasal cannula therapy | 4.8 | 1.6 |
| 6 | Hospitalised, requiring invasive mechanical ventilation ± additional organ support | 4.0 | 1.6 |
| 7 | Death | 10.7 | 12.6 |

Table 1 also shows the percentage of participants expected to be in each state at Days 14 and 28 under standard care (empirical estimate, based on a small convenience sample of accessible study results available at the time of writing). The secondary and exploratory outcomes are listed in Table 2.

**Table 2:** Secondary and exploratory outcomes for the CLARITY trial. ^*^AKI is defined as any documentation of the following: increase in serum creatinine by ≥ 0.3 mg/dl (≥ 26.5 *µ*mol/l) within 48 hours; increase in serum creatinine to ≥1.5 times baseline, which is known or presumed to have occurred within the prior 7 days; or urine volume <0.5 ml/kg/h for 6 hours [1].

| Outcome | Additional information |
| --- | --- |
| **Secondary:** |  |
| 7-point ordinal scale score | As at Day 28 |
| All cause mortality | As at Day 28<br>As at Day 90 |
| Admission to ICU | Time to admission between Day 0 and 28<br>Time to admission between Day 0 and 90<br>Number of ICU free days between Day 0 and 90 |
| Respiratory failure | Time to non-invasive or invasive mechanical ventilation between Day 0 and 28<br>Number of ventilator-free days between Day 0 and 28 |
| Kidney failure | Time to requirement for dialysis between Day 0 and 28<br>Number of dialysis free days between Day 0 and 28 |
| Hospitalisation (length of stay) | Number of hospitalisation days between Day 0 and 28<br>Number of hospitalisation days between Day 0 and 90 |
| Acute Kidney Injury (AKI)* | Time to AKI between Day 0 and 28 |
| Hypotension requiring vasopressors | Time to hypotension requiring vasopressors between Day 0 and 28 |
| **Exploratory:** |  |
| Hyperkalemia | Time to hyperkalemia between Day 0 and 28 (any episode of serum potassium > 6.0mmol/L) |
| Oxygen requirement | Oxygen support free days between Day 0 and 28 |

ARBs have been extensively researched and used widely in standard practice for around thirty years, forming a significantly robust safety profile. The trial endpoints capture the commonly known safety events associated with ARBs. No additional safety reporting of adverse events or serious adverse events is required given the well-understood safety profile of these agents.

### 2.7 Data management

The Data Management Plan for CLARITY has been developed according to the standard operating procedures of the NHMRC Clinical Trials Centre at the University of Sydney, and the Trial Master File is held centrally by the Clinical Trials Centre. All such documentation are currently not publicly available.

## 3 Statistical analysis framework

All outcomes will be analysed within a Bayesian framework based on joint posterior distributions. Sampling from the posterior distribution will use Markov chain Monte Carlo (MCMC) methods [21, 12, 11], and where necessary, model choice will be determined via an appropriate information criterion [31]. Convergence of MCMC chains will be assessed via trace plots and the Gelman-Rubin diagnostic [9], and goodness-of-fit will be assessed via posterior predictive checks. All computation will be performed within the R-package which will interface with STAN [3] or equivalent should the need arise.

Across all analyses, weakly informative prior information calibrated by prior predictive checks will be used such that trial conclusions will be predominately data-driven. For all parameters of interest, posterior means will be reported along with 95% credible intervals and the posterior probability that the parameter is greater or less than 0.

### 3.1 Descriptive summaries

#### 3.1.1 Baseline characteristics

Baseline characteristics will be summarised for each treatment arm and overall. Discrete variables will be summarised by frequencies and percentages. Percentages will be calculated according to the number of participants for whom data are available. Where values are missing, the denominator (which will be less than the number of participants assigned to the treatment group under consideration) will be stated. Continuous variables will be summarised by use of standard measures of central tendency and dispersion using mean and standard deviation and/or median and first and third quartiles. Free text entries for fields collecting both categorical and free text information (e.g. ethnicity) will be assessed and assigned to a category if appropriate. No testing will be performed for differences in baseline characteristics between treatment arms as per CONSORT principles.

Table 3 shows the baseline characteristics that will be summarised.

**Table 3:** Baseline characteristics

| Demographics |  |
| --- | --- |
|  | Sex<br>Age<br>Ethnicity |
| Medical History |  |
| COVID-19 history and Management | Intended for hospital or for community management |
| Co-morbidities | Chronic Kidney Disease<br>Hypertension<br>Diabetes<br>Cardiovascular disease (including heart failure, ischemic heart disease, acute myocardial infarction, congenital heart disease, stroke, peripheral vascular disease)<br>Cancer in last 5 years (not including basal cell carcinoma and squamous cell carcinoma)<br>Chronic respiratory illness<br>Severe Liver Disease (Child-Pugh-Turcotte score 10-15 or biliary obstruction)<br>Other relevant medical conditions<br>Pregnancy status (pregnant or breastfeeding) |
| Medications | Renin-Angiotensin-Aldosterone System (RAAS) inhibitor (including ACEi, ARB, Aldosterone antagonist, Angiotensin Receptor-Neprilysin Inhibitor, Aliskiren)<br>Non-RAAS inhibitor Blood Pressure (BP) lowering agent<br>Glucose lowering medication<br>Steroids<br>Steroid inhalers/nasal spray<br>COVID-19 specific therapies<br>Other immunosuppressants<br>Other inhalers<br>Vasopressor<br>Proton-pump inhibitor (e.g. omeprazole, esomeprazole, pantoprazole)<br>Aspirin<br>Lipid lowering agent (e.g. statins, fibrates)<br>Antimicrobials<br>Other |
| Smoking status | Current, previous, non-smoker |
| Physical Examination |  |
|  | Blood pressure (mmHg)<br>Height (cm)<br>Weight (kg)<br>Body mass index (kg/m<sup>2</sup>) |
| Laboratory Measures |  |
|  | Serum creatinine (mg/dL)<br>Estimated Glomerular Filtration Rate (mL/min/1.73 m<sup>2</sup>)<br>White Cell Count (x10<sup>9</sup>/L)<br>Neutrophils and Lymphocytes (x10<sup>9</sup>/L)<br>D-Dimer (mg/L FEU or mg/L DDU)<br>C-Reactive Protein (mg/L or nmol/L)<br>Creatine Kinase (U/L) |

#### 3.1.2 Delivered treatment

Medication adherence will be reported by each treatment arm as adherence days (median, first and third quartiles, minimum and maximum), number of days of follow-up (median) and percentage of days medication was administered (mean ± SD) for those who complete follow-up and those who do not. We will also report the mean average dose of treatment for participants randomised to ARB plus standard care, the use of other COVID-19 directed care, other hypertensives and use of oxygen.

### 3.2 Analysis of primary outcome

The primary outcome for this trial is a 7-point ordinal scale of clinical outcomes (Table 1), assessed on Day 14 and will be modelled using proportional odds cumulative logistic regression [20]. Under such a proportional odds model, the treatment effect (along with the parameters associated with other variables) are common among the response categories.

The model is specified as follows. Let *Y*_*i*_ ∈ { 1, …, *K*} denote a random variable for the outcome for the *i*th participant where the ordering is natural and *K* = 7. A proportional odds model can be constructed based on the categorisation of a latent continuous variable *Y* ^*^. Ordered cut-points *c* ∈ ℝ^*K*−1^ are defined such that *c*_*k*_ *< c*_*k*+1_ for *k* ∈ {1, …, *K* − 2} then for *k* ∈ {1, …, *K*}:

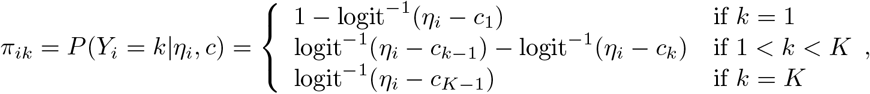

where *Y*_*i*_ ∼ *MN* (*n* = 1, (*π*_*i*1_, …, *π*_*i*7_)) and ‘*MN* ‘ denotes the multinomial distribution. The linear predictor 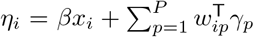 comprises a parameter *β* that charaterises the treatment effect, with *x*_*i*_ = 1 for ARB treatment group membership and *x*_*i*_ = 0 otherwise and a vector of parameters *γ* for modelling variation in the response associated with baseline variables defined in 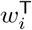. Note that this implementation will be such that exp(*β*) *<* 1 results in a reduction in the probability of death.

The model is parameterised with the cuts *c*_*k*_ derived from a simplex with a Dirichlet prior and concentration of 1. All fixed effect terms will use weakly informative normal priors centred on zero with a standard deviation of *σ*_*β*_ = 10. If issues with convergence or sampling arise, variations to these priors are permissible at the discretion of the analyst, however, all model variations and their justification will be reported to the DSMB and will be disclosed in all internal and external publications.

The motivation to adjust the model for baseline covariates stems from the understanding that mortality rates associated with COVID-19 infection vary by age, sex, ethnicity and the presence of some comorbid diseases [19, 34, 25]. Specifically, the case mortality rate is estimated as being very low (0.06% or less) in age groups under 50, 0.5% in 50-59 year olds, 2.9% in 60-69 year olds before steeply rising to 40% in those aged 90 and over [19]. Additionally, disease severity appears to be worse in males, in those with pre-existing comorbidities, such as hypertension, diabetes, heart failure, chronic kidney disease, and chronic respiratory illness [34], and in the presence of obesity [25].

Since age, sex, co-morbid disease and oxygen requirement are the most supported, baseline values for these variables will be included in the linear predictor as fixed effects with levels as summarised in Table 4. Incorporating these terms in the model allows us to account for potential baseline imbalance, obtain a stratified estimate of the response and can increase power. All decisions arising from successive planned analyses will be based on the adjusted model, however, both unadjusted and adjusted results will be reported.

**Table 4:**
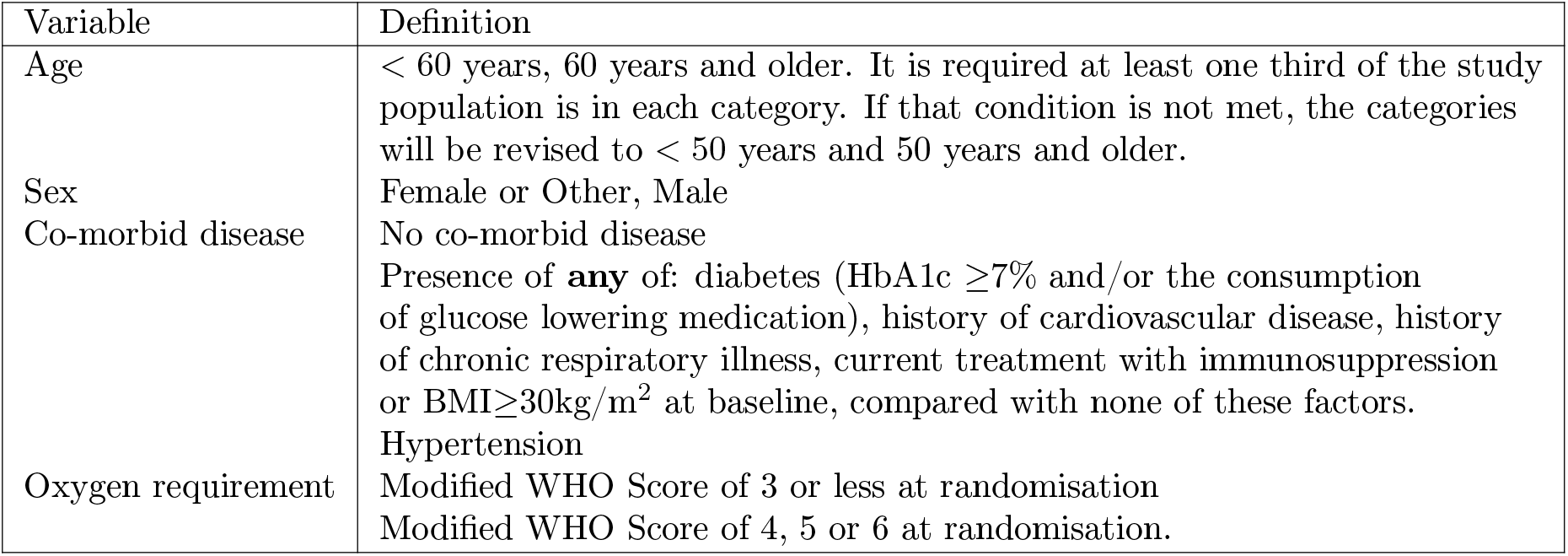
Prognostic baseline characteristics/pre-specified subgroups for analyses of the primary, secondary and exploratory outcomes.

#### 3.2.1 Sensitivity to assumptions

Sensitivity analyses for the primary outcome will be undertaken to determine how the results of the analysis may change depending upon particular modelling assumptions. For this trial, this will include:

- choice of priors
- relaxing the proportional odds assumption

Additionally, a subset of data will be modelled constrained to participants who were randomised to receive either an ARB or a placebo (i.e. those participants randomised in India). Technically, this is a subgroup analysis, but is included here because the results from this subset of the data represent an important consideration for interpreting the results for the primary outcome.

Weakly informative priors have been pragmatically chosen to regularise estimates and minimise problems with posterior sampling. The sensitivity analysis will evaluate the impact of uninformative, skeptical and enthusiastic priors [2], keeping in mind the comments of [10] on the influence of the likelihood on interpreting the contribution of the prior.

Relaxing the proportional odds assumption accommodates category specific effects. For example, an intervention could be potentially useful for healthier participants, but harmful to participants that are already very sick. The cumulative ordinal model is not well suited for this purpose (category specific effects in this model can lead to negative probabilities of having some scores), so an alternative modelling strategy is required. One approach is to collapse the response into several binary comparisons, fit separate logistic regression models (response scores 1 versus 2-7, 1-2 versus 3-7 etc) and qualitatively compare the odds ratios obtained from these models to the more parsimonious proportional odds model. However, this becomes unwieldy for a large number of response categories. Therefore, the sensitivity analysis will be undertaken using an adjacent category model with category-specific effects to gain insight into heterogeneity in the treatment effect. Metrics for relative goodness-of-fit will be obtained via leave-one-out cross validation.

The category specific effects model will only be evaluated at the final analysis, but the potential for violations in the proportional odds assumption will be monitored heuristically at each planned analysis.

### 3.3 Analysis of secondary and exploratory outcomes

The analysis of all secondary and exploratory outcomes detailed in Table 2 will be model-based within a Bayesian inference framework. An outline of the modelling approach for each of the different data types are given below. Unless directed by the DSMB, the analysis of all secondary and exploratory outcomes will only be conducted as part of the final analysis.

#### 3.3.1 Binary outcome

The following outcome is represented as a binary random variable, and will be summarised by number of and proportion of outcomes within each category. The relative odds of the outcome will be modelled by treatment assignment using standard logistic regression.

- (All-cause) mortality at Day 28 (participants discharged prior to Day 28 will be considered alive unless noted otherwise)

For logistic regression, the parameter associated with the treatment term in the model reflects the change in the log-odds of the event relative to the log-odds of the event in the reference group holding all other terms constant.

#### 3.3.2 Ordinal outcomes

The following outcomes can be represented as ordinal scales. Each will be summarised by the proportion in each category, and modelled using the same approach described for the primary outcome. Specifically, the relative odds of the outcome will be modelled by treatment assignment using proportional odds logistic regression.

- 7-point ordinal scale score at Day 28 (primary is at Day 14)
- Number of ventilator-free days between Day 0 and 28
- Number of oxygen support free days between Day 0 and 28
- Number of dialysis-free days between Day 0 and 28
- Number of ICU-free days between Day 0 and 90

For the latter four outcomes, the occurrence of death will be coded such that it is the worst possible outcome, with the remaining possible outcomes being coded via the natural ordering.

The interpretation of the parameter associated with treatment term in the proportional odds cumulative ordinal model is very similar to that of a logistic regression. For the ordinal model, one can conceptualise the event as successive splits of the scores demarcating success. So, for example, one might consider realising 1 or more ventilator-free days (VFDs) versus 0 or worse. Equivalently, one can consider realising 2 or more VFDs versus 1 or worse and so on. The parameter estimate associated with treatment simply characterises the change in the log-odds of the event (as defined above) on ARB relative to the log-odds of the event on placebo, holding all other terms constant. The proportional odds assumption holds that the transitions between each of the splits are equally impacted by the treatment, i.e. there are no category-specific effects.

#### 3.3.3 Time-to-event outcomes

The following outcomes are considered in terms of the time until the occurrence of an event, all of which occur in the presence of a competing risk. For example, time to admission to ICU is blocked should the patient die prior to being admitted to ICU. The goal is to characterise the rate or intensity of events by treatment assignment.

- Time to discharge alive from hospital between Day 0 and 28
- Time to discharge alive from hospital between Day 0 and 90
- Time to admission to ICU between Day 0 and 28
- Time to admission to ICU between Day 0 and 90
- Time to non-invasive or invasive mechanical ventilation between Day 0 and 28
- Time to requirement for dialysis between Day 0 and 28
- Time to AKI between Day 0 and 28
- Time to hypotension requiring vasopressors between Day 0 and 28
- Time to hyperkalemia between Day 0 to 28

With the exception of time to discharge (where time to discharge is the event of interest) each of these events exist in the presence of the competing risk of death and/or discharge. Standard approaches for evaluating competing risk include models that focus on the cause-specific hazard and/or the subdistributional hazard [26, 7]. The former is preferred where interest is in the rate of occurrence rather than the risk and is generally recommended for evaluating direct causal effects. Therefore, we will adopt a piecewise exponential model for the cause-specific hazards (reporting results from all event types). The linear predictor will be of a similar form as described in Section 3.2; the exponential of which will estimate the multiplicative shift away from the baseline marginal hazard function.

The cause-specific hazard is the instantaneous risk of failure from a specific cause given that no failure from any cause has happened. Interpretation of the parameter estimate associated with treatment effect will be in terms of the effect on the cause-specific hazard of the event under consideration. Finally, in a competing risk setting, the cumulative incidence function (a generalisation of one minus the survival to the competing risk setting) cannot be estimated naively using one minus the Kaplan-Meier estimator. Therefore, any visualisation of the survival and cumulative incidence will account for this.

### 3.4 Subgroup analyses

Rather than split the data into strata and proceed with independent analyses, subgroup analyses will be undertaken by extending the linear predictor in the specified models to account for treatment by group interactions. The pre-specified subgroups include age, sex, co-morbid disease and oxygen requirement at baseline as detailed earlier in Table 4.

Shrinkage through prior specification will be used as a method for mitigating issues associated with multiplicity. Additionally, only first-order interactions will be considered, which implicitly assumes all higher order interactions are set to zero. Specifically, for the subgroup treatment effects, a revised linear predictor will be introduced from the earlier specification in Section 2.6:

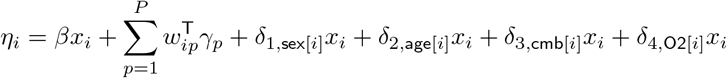

where each *δ*_*g*_ parameter corresponds to a vector of parameters associated with each subgroup of interest with *δ*_*g*_ ∼ Normal(0, *σ*_*δ*_) and *σ*_*δ*_ ∼ Exponential(1) such that the interaction terms are estimated under the assumption of a common variance [6]. The abbreviations “cmb” and “O2” correspond to “comorbidity” and “oxygen requirement”, respectively.

From the above model, each covariate can be considered separately and standardised treatment effects obtained in each subgroup as weighted averages. For example, to obtain a standardised treatment effect for males, compute

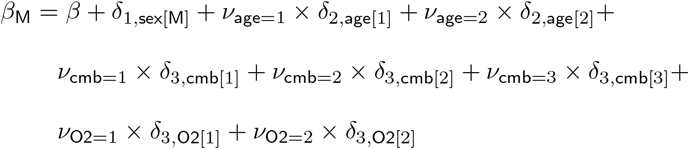

where *β*_M_ denotes the log odds-ratio for the treatment effect in males, *ν*_age=1_ denotes the observed relative frequency of males in the first age category, *ν*_age=2_ denotes the observed relative frequency of males in the second age category and the other *ν* weights are defined similarly.

### 3.5 Missing data

Among other possibilities, missing data are to be anticipated due to loss to follow-up and competing priorities on institutional resources to complete follow-up.

Under the missing completely at random (MCAR) and missing at random (MAR) assumptions, there will be loss of precision due to the missingness, but there is no bias in the parameter estimates when appropriate statistical methods are used. However, under missing not at random (MNAR) the probability of missingness depends on the (unobserved) missing values. When missingness is due to MNAR, there is both a loss in precision and bias, and therefore sensitivity analyses are required. MAR is commonly assumed, although MNAR is arguably more applicable for most settings. In the CLARITY trial, there is potential for MNAR because differential loss to follow-up could be observed across treatment groups, however this is believed to be unlikely.

Patterns of missingness for the outcome and covariates will be reported by time and group. When applicable, missing outcome data will be imputed using the standard posterior predictive approaches within Bayesian inference [28]. The posterior distribution for imputation will be based on the analysis of all available data. Missing covariate information may also be imputed, and this will be based on, where possible, other available data (e.g. missing region or site).

If concerns arise regarding missingness (or a sensitivity analysis is requested by the DSMB), then single value imputation may be used implementing a best-worst-case and worst-best-case sensitivity analyses to evaluate the potential range of impact of missing values.

### 3.6 Exploratory analyses

#### Temporal treatment effect heterogeneity

As there is potential for changes in standard of care and variation in the circulating virus strains, we will explore temporal heterogeneity in the treatment effect for the primary outcome. To do so, indicator variables will be created that categorise trial epochs. These indicator variables will then be included (as described in Section 3.2) as main effects and as terms that interact with treatment. Choice of the time step will be pragmatically selected based on obtaining reasonably sized groups for the purposes of estimation.

#### Disease progression modelling

Given the potential for participants to manifest complex disease progression, state-space modelling will be undertaken to characterise transition probabilities and sojourn times.

### 3.7 Planned analyses

The first planned analysis will be conducted as soon as practicable after the 700th enrolled participant has been followed for 14 days. This was decided by exploring the sampling distribution of the treatment effect parameter for a variety of different scenarios (Section 4) including different odds ratios, and a desire to have a relatively stable estimate, see Figure 2. From the results, the first planned analysis at 14 days after 700 participants were enrolled was a reasonable compromise. After this, the planned analyses will occur 14 days after every 300 additional participants have been enrolled. The decision also considers practical constraints such as needing enough time to complete the analyses and having these reviewed by the DSMB before the next planned analyses.

**Figure 2:**
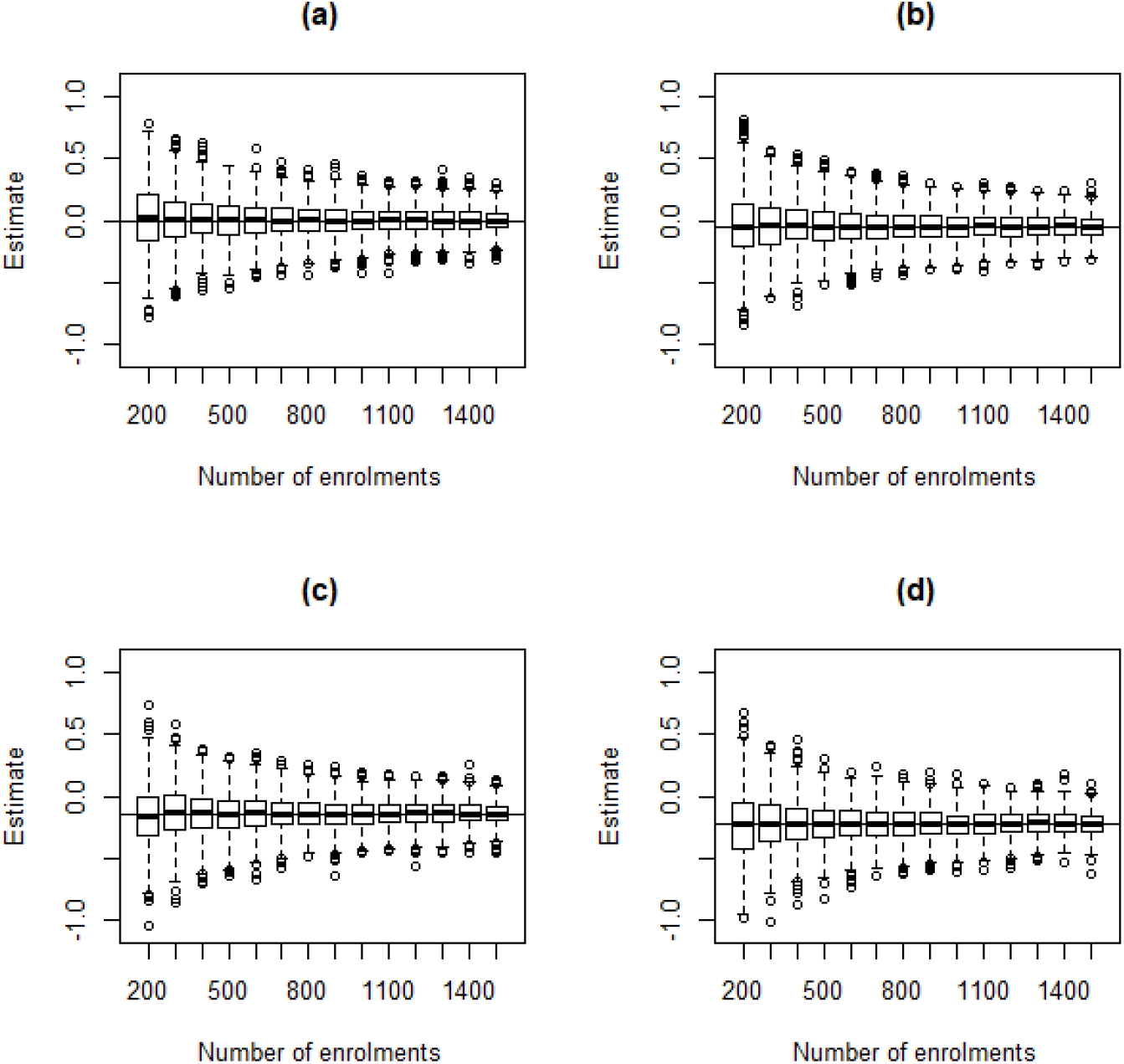
Distribution of the posterior mean of *β* based on 1000 simulated data sets for a given number of enrolments and odds ratios of (a) 1.00, (b) 0.95, (c) 0.87 and (d) 0.80.

The planned analyses will model the primary outcome (Section 3.2) for which both the unadjusted (i.e. model only includes treatment status) and adjusted posterior treatment effects will be reported to the DSMB. Decision rules will be evaluated based on the results from the adjusted primary analysis.

Subgroup analyses (Section 3.4) will only be undertaken at the final analysis as these are essentially exploratory in nature and need to be interpreted with great care.

Based on the results from the primary analysis, supplemented by external knowledge, recommendations will be made to the Trial Steering Committee by the DSMB to either halt or continue recruiting into the trial.

### 3.8 Decision rules

Pre-specified decision rules will be evaluated at each planned analysis, and the trial can be stopped based on the results of these analyses. This implies that the trial sample size is a random variable with an upper bound of 2200, dictated by the available resources. For this trial, two decision rules were adopted. These were to assess treatment effectiveness, and to determine if it is futile to continue the trial. Both of these decision rules are based on the primary outcome as described in Section 3.2 accounting for differences between treatments arms and baseline characteristics only. In the following section, each decision rule is defined, including the methods by which each will be evaluated.

#### 3.8.1 Effectiveness

At each planned analysis, the predictive probability of treatment arm effectiveness compared to placebo or standard care alone (Australia only has standard care) will be assessed. To do so, one may consider that there are two types of participants in the interim data; (1) participants who have been enrolled and had their Day 14 outcome ascertained; and (2) participants who have been enrolled but have not yet had their Day 14 outcome ascertained. The expectation of the probability that the posterior probability *β <* 0 is greater than the pre-specified level of evidence (Δ = 0.975) will be evaluated, which is nominally consistent with a standard one-sided frequentist type-I error of 0.025. The expectation is taken over the data from participants who have been enrolled but have not responded and is assessed against a decision threshold, *δ*_*e*_ set at 0.95. Specifically, the expectation is defined:

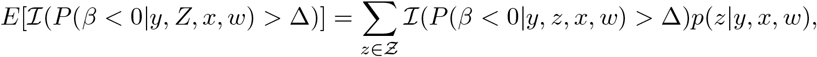

where ℐ() is an indicator function which equals one if the event is true and zero otherwise, *y* denotes data on the primary outcome for the participants who have been enrolled and had their Day 14 outcome ascertained, *x* and *w* denote treatment allocations and baseline characteristics (respectively) for all participants enrolled, and *Z* ∈ *Ƶ* is the random variable associated with *z* which denotes supposed future data for the participants who have been enrolled but not yet had their Day 14 outcome ascertained.

Evaluating the above expectation is unwieldy, so simulation will be used to form an estimate. The approach for this is outlined in Algorithm 1 where the data *y*, the treatment allocations *x*, baseline characteristics *w* and the prior on the parameters *p*(*β, c*) are initialised. For large *B*, a sample from the posterior distribution of the parameters is obtained based on data from participants who have been enrolled and have had their Day 14 outcome ascertained. For those participants who have not yet had their Day 14 outcome ascertained, their treatment allocation and baseline characteristics are known but their outcome is not, so their outcome is simulated (line 4). This forms a (partly simulated) data set for all participants enrolled in the trial from which a posterior distribution can be found.

##### Algorithm 1 Estimation of decision rule for treatment effectiveness

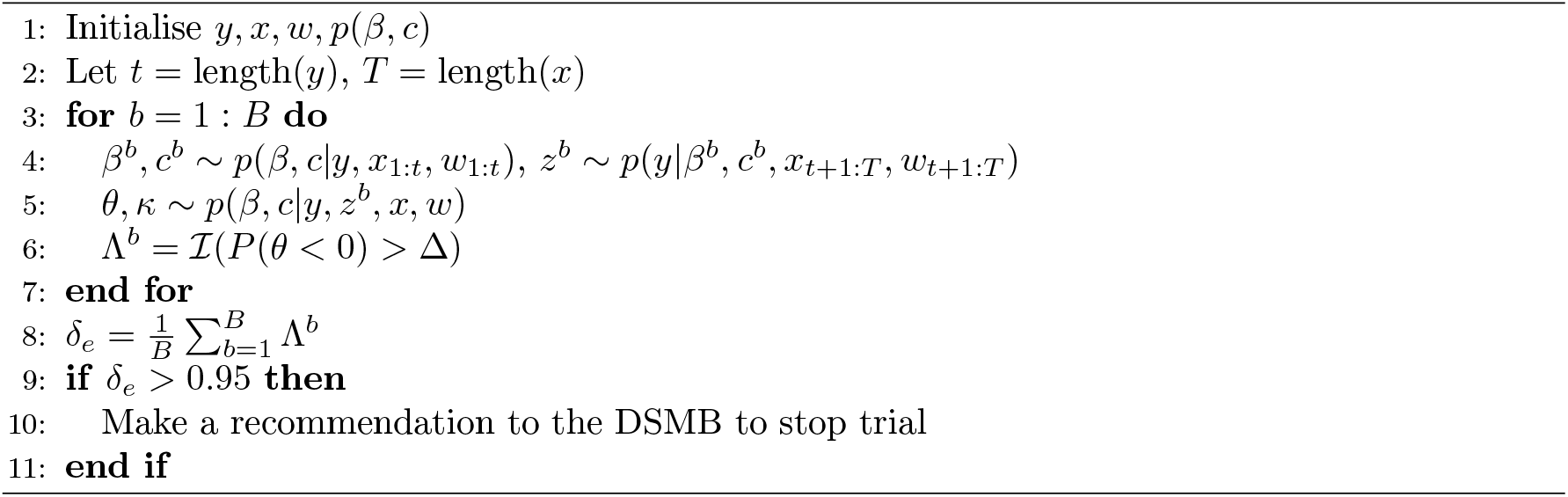

To sample from this posterior distribution, MCMC could be used. However, given this step needs to be performed a large number of times (within a single simulation of a trial and across different trial settings), using MCMC here becomes computationally infeasible. Accordingly, a fast approach to form an approximation of the posterior distribution is needed. Consequently, the Laplace approximation [17] will be adopted, as has been used previously for a similar purpose [23, 30]. Based on this posterior, an indicator function is evaluated to determine whether treatment effectiveness would be concluded at the pre-specified level of evidence (Δ = 0.975), if the outcomes from all enrolled participants were ascertained (line 6).

After repeating this procedure a large number of times, the probability the trial is expected to be successful is approximated (line 8). If this is larger than a nominal value of *δ*_*e*_ = 0.95, then the trial will stop for expected treatment effectiveness.

#### 3.8.2 Futility

The purpose of the futility decision rule is to determine whether it would be futile to continue the trial based on the apparent likelihood of concluding effectiveness. To evaluate this, we consider the probability of concluding treatment effectiveness if the trial were to continue until the maximum sample size of *N* = 2200. At each planned analysis there will be three types of participants: (1) participants who have been enrolled and had their Day 14 outcome ascertained; (2) participants who have been enrolled but have not had their Day 14 outcome ascertained; and (3) participants who have not yet enrolled and therefore could not have had their Day 14 outcome ascertained.

As in the effectiveness decision rule, there will be uncertainty about the responses for a number of participants. Given this, we consider the expectation of an indicator of success over the distribution of these unknowns. Specifically, the expectation is defined:

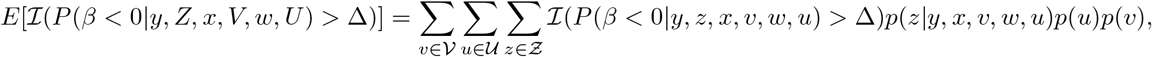

where *V, U* and *Z* are the random variables associated with *v* which denotes supposed future treatment allocations, *u* which denotes future baseline characteristics of participants and *z* which denotes supposed future outcome data, respectively. The distribution of *V* denoted as *p*(*v*), will be based on the 1:1 randomisation and the distribution of *U* denoted as *p*(*u*), will be estimated non-parameterically via re-sampling *w* (independently and with replacement).

##### Algorithm 2 Estimation of decision rule for futility

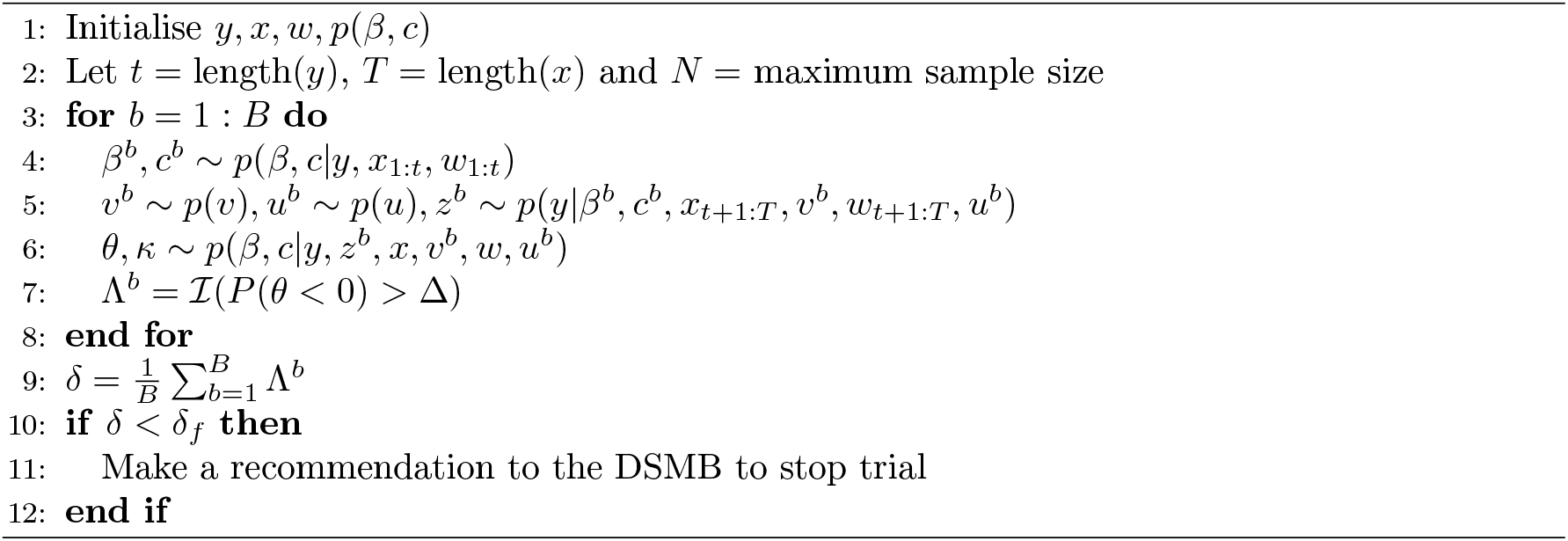

Simulation is again used to form an estimate, and the approach for this is outlined in Algorithm 2 where *y, x, w* and *p*(*β, c*) are initialised. For large *B*, a sample from the posterior distribution for the parameters based on data from participants who have responded is drawn (line 4). The remaining trial data is then simulated (line 5). For this, some treatment allocations and baseline characteristics are known and some are not. For those that are not known, these are simulated from *p*(*v*) and *p*(*u*) as noted above. Data for the remainder of the trial (up to the maximum sample size of *N* = 2200) are then simulated (again in line 5) and a sample from the posterior distribution of the parameters based on this partly simulated data set is then obtained (line 6). Based on this posterior, an indicator function is evaluated to determine whether treatment effectiveness would be concluded at the pre-specified level of evidence (Δ = 0.975), if the outcomes from all enrolled participants were ascertained (line 7).

After repeating this procedure a large number of times, the probability the trial will conclude that the intervention is effective is approximated (line 9). If this is less than a pre-specified value *δ*_*f*_ = 0.02 (suggesting that concluding treatment effectiveness after *N* enrolments is very unlikely), then it will be concluded that it is futile to continue with the trial.

## 4 Specification of tuning parameters for decision rules

The specification of the two decision rules adopted in this trial requires pre-specifying the values of Δ, *δ*_*e*_ and *δ*_*f*_. To do so, the statistical power and type-I error of the trial were explored through simulation, aiming to find appropriate values for these parameters. To run such a simulation study, a number of assumptions needed to be made. For the simulations for this trial, the following was assumed:

- The data observed in the trial will follow a proportional odds logistic regression model where any average difference in outcomes can only be explained by differences in their assigned treatments;
- The data observed from each participant in the trial will be independent. That is, each participant yields a single observation from the 7-point ordinal scale, and these are independent between participants;
- Five values for the treatment effect were explored. These relate to odds ratio values (i.e. exp(*β*)) of 1.0, 0.95, 0.87, 0.8 and 0.67;
- Accrual rate of participants into the trial will be either 120, 100 or 80 per month. That is, three potential scenarios which can be defined as ‘best’, ‘expected’ and ‘worst’ (respectively) from the trial perspective;
- The probability of observing each category of the 7-point ordinal scale for a participant within the placebo plus standard care group is as shown in Table 5. Here, three potential scenarios (‘best’, ‘expected’ and ‘worst’ from the patient perspective) have been defined;
- Planned analyses will not start until 14 days after 700 participants have enrolled into the trial. After this, planned analyses will occur 14 days after an additional 300 participants have been enrolled in the study.

**Table 5:** Expected distribution (as percentages) of 7-point ordinal scale at Day 14 for placebo/Standard of care group where ‘best’, ‘expected’ and ‘worst’ case is from the patient’s perspective.

| Categories | Best | Expected | Worst |
| --- | --- | --- | --- |
| Not hospitalised, no limitations on activities | 16.0 | 0.1 | 0.1 |
| Not hospitalised, some limitation on activities | 28.6 | 31.0 | 19.4 |
| Hospitalised, not requiring supplemental oxygen | 32.0 | 34.0 | 30.0 |
| Hospitalised, requiring supplemental oxygen | 13.0 | 15.5 | 20.0 |
| Hospitalised, requiring non-invasive mechanical ventilation or high flow nasal cannula therapy | 2.4 | 4.7 | 7.0 |
| Hospitalised, requiring invasive mechanical ventilation ± additional organ support | 2.0 | 4.0 | 5.5 |
| Death | 6.0 | 10.7 | 18.0 |

Based on these assumptions, 500 trials were simulated for every combination of different scenarios as outlined above. That is, trials were simulated with five different odds ratios, three different accrual rates and three different expected outcomes in the control group. This means for each odds ratio there are nine different scenarios, and it is of interest to determine the chance of stopping due to declaring effectiveness or futility across these settings, and also how variable trial outcomes are across these different scenarios. To evaluate expected effectiveness and futility, data were simulated 500 times (i.e. *B* = 500 in Algorithms 1 and 2).

After extensive exploration, it was determined that Δ = 0.975, *δ*_*e*_ = 0.95 and *δ*_*f*_ = 0.02 yielded acceptable results across a variety of scenarios. Figure 3 provides a summary where (a) shows the probability of concluding treatment success at the end of the trial, (b) shows the probability of stopping the trial early due to futility, and (c) shows the average sample size where this average is taken across all 500 trials regardless of whether it was stopped early or no conclusion was drawn.

**Figure 3:**
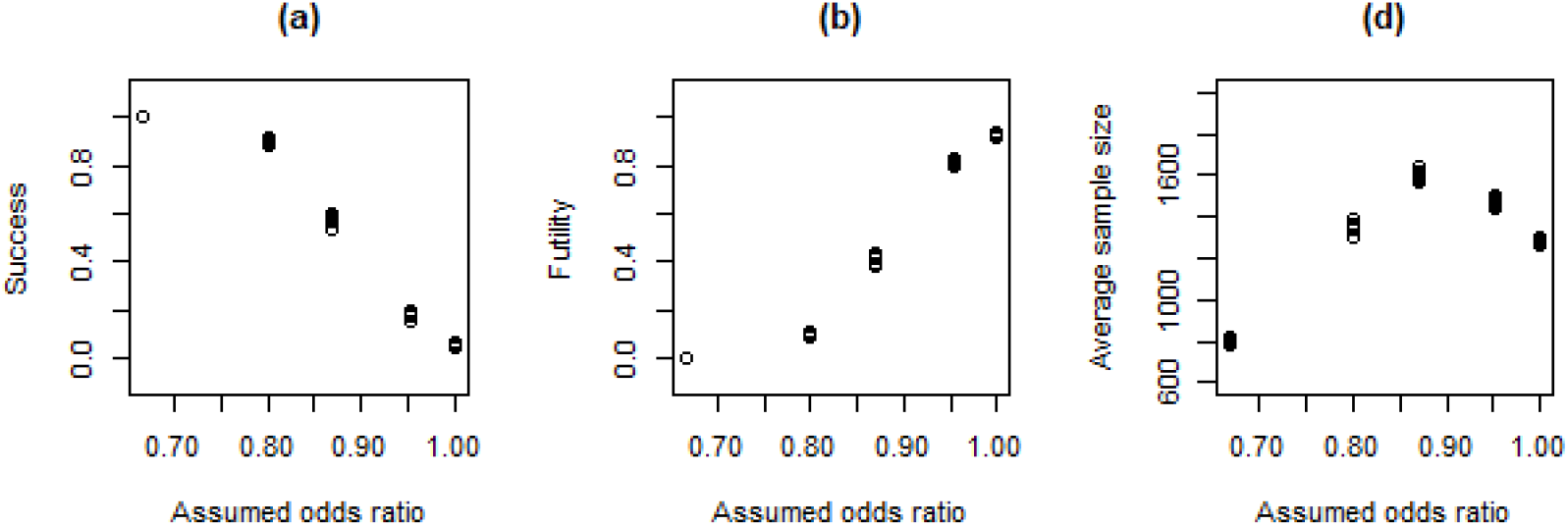
Estimated probability of the trial stopping for (a) Success, (b) Futility, and (c) average sample sizes based on a variety of different assumptions about the odds ratio, recruitment rate and data that will be observed.

We note that for each odds ratio, there are nine scenarios, so each plot actually shows nine points for a given odds ratio. Some of these points are difficult to see because there is very little variability in outcomes across the nine scenarios. This is reassuring as it means the outcomes of the proposed design appear robust to foreseen variations in accrual rate and expected outcomes within the control group. As would be expected, there is substantial variability between different assumed odds ratios. In particular, when the assumed odds ratio is 0.8 and below, the power of the trial to determine that the intervention is effective remains above 80%. When the assumed odds ratio is 0.87, the power reduces to around 60%, and there is very little power when the assumed odds ratio is 0.95. When there is no benefit on treatment, the type-I error remains around 5%.

The average sample size shows that the maximum average is less than 1,650 and this occurred when the assumed odds ratio was 0.87. This decreases as the assumed odds ratio decreases because the effectiveness decision rule is, on average, met earlier. As the assumed odds ratio increases, the average sample size also decreases because the futility rule is stopping those simulations early. Given the uncertainty of the clinical importance of a benefit associated with an odds ratio of 0.95, this appears reasonable. For comparison, the required sample size was also computed for a variety of odds ratios and levels of statistical power based on a standard randomised controlled trial where there are two treatment arms and the outcome is ordinal. For this, the analytic solution for sample size as given by [32] was used. These results are shown in Table 6, demonstrating that larger sample sizes are needed to achieve equivalent statistical power to the adaptive trial. For example, to achieve a power of 60% for an odds ratio of 0.87, 2,391 participants need to be enrolled. This is compared to an average of 1,650 in a trial with an adaptive sample size. This highlights one of the potential benefits of adopting an adaptive sample size approach.

**Table 6:** Required sample size for a standard, two-arm, randomised controlled trial for different odds ratios and levels of statistical power.

|  |  | Odds ratio |  |  |  |  |  |  |  |
| --- | --- | --- | --- | --- | --- | --- | --- | --- | --- |
|  |  | 0.95 | 0.91 | 0.87 | 0.83 | 0.8 | 0.77 | 0.74 | 0.71 |
| Power | 0.50 | 14732 | 3861 | 1796 | 1055 | 705 | 510 | 390 | 310 |
|  | 0.55 | 17069 | 4473 | 2081 | 1223 | 817 | 591 | 452 | 359 |
|  | 0.60 | 19620 | 5142 | 2391 | 1406 | 938 | 679 | 519 | 413 |
|  | 0.65 | 22443 | 5882 | 2736 | 1608 | 1073 | 777 | 594 | 472 |
|  | 0.70 | 25623 | 6715 | 3123 | 1835 | 1225 | 887 | 678 | 539 |
|  | 0.75 | 29292 | 7676 | 3570 | 2098 | 1401 | 1013 | 775 | 616 |
|  | 0.80 | 33665 | 8822 | 4103 | 2411 | 1610 | 1165 | 890 | 708 |
|  | 0.85 | 39147 | 10259 | 4771 | 2804 | 1872 | 1354 | 1035 | 824 |
|  | 0.90 | 46631 | 12220 | 5683 | 3340 | 2230 | 1613 | 1233 | 981 |

Overall the rules and design of this trial appear to achieve reasonable operating characteristics for moderate to large odds ratios across a variety of scenarios that might be observed. It should be noted that a variety of additional scenarios were also considered in terms of different values for *δ*_*e*_ and *δ*_*f*_. Across all of these scenarios, the values proposed here appear to yield more desirable operating characteristics, given the trial settings and the assumptions made. In addition, alternative primary outcomes such as the 7-point ordinal scale ascertained on Day 28, a binary outcome being some combination of the 7-point ordinal scale ascertained on Day 14 and Day 28, and time-to-ventilation/death were also explored to determine whether a reduced sample size could be considered for this trial. However, no significant improvement in efficiency was observed. Details of this for the time-to-event outcome are given in the appendix.

## 5 Discussion

The CLARITY trial investigates the effectiveness of ARBs with standard care compared to placebo (if provided) with standard care to reduce the duration and severity of COVID-19 in severe participants.

The first patient was enrolled on 18th August, 2020. Currently there are 14 and 7 active sites in India and Australia, respectively. Recruitment has been predominately from India reflecting the relative caseload during the period the trial has been active. Due to uncertainty about the effectiveness of ARBs for COVID-19 and the urgent need to find effective treatments for COVID-19 patients, the trial was designed with an adaptive sample size. The decision rules were constructed to maintain desirable levels of statistical power and type-I error properties, while maintaining a relatively small average sample size, particularly when compared to a traditional randomised controlled trial. Other primary outcomes were explored (including a binary and a time-to-event outcome), however negligible gain in efficiency was observed.

In terms of the decision rules, the trial can be stopped due to sufficient evidence of effectiveness or futility. It is worth noting that an additional decision rule related to harm was also explored. That is, a rule that would stop the trial if there was sufficient evidence that the treatment plus standard care arm was performing worse than the placebo plus standard care arm (or less than a minimum important clinical difference). However, little to no benefit in terms of the trial operating characteristics was observed when this decision rule was included. This appeared to be because whenever this rule was triggered, the futility rule was also being triggered. Thus, it was concluded that this additional decision rule was not required.

### Trial status

The trial commenced recruitment on 18th August, 2020 and the first planned analysis is now imminent.

## Data Availability

The data are currently being collected as part of an active clinical trial.

## 6 Supplementary information

## 6.1 List of abbreviations

ARBs: Angiotensin Receptor Blockers;
AT1R: Angiotensin II type 1 receptor;
BERN: Bernoulli;
CONSORT: Consolidated Standards of Reporting Trials;
CLARITY: Controlled evaLuation of Angiotensin Receptor Blockers for COVID-19 respIraTorY Disease;
COVID-19: Coronavirus disease 2019;
DSMB: Data Safety Monitoring Board;
ICU: Intensive care unit;
MAR: Missing at random;
MCAR: Missing completely at random;
MCMC: Markov chain Monte Carlo;
MNAR: Missing not at random;
RAAS: Renin-Angiotensin-Aldosterone System;
RAS: Renin-angiotensin system;
SAP: Statistical analysis plan;
SARS-CoV-2: Severe acute respiratory syndrome coronavirus 2;
SD: Standard deviation;
VFDs: Ventilator-free days

## 6.2 Ethics approval and consent to participate

The CLARITY trial is approved by the Sydney Local Health District Ethics Review Committee (Royal Prince Alfred Hospital Zone; Code: EC0113) in Australia (X20-0118 & 2020/ETH00742) and The George Institute for Global Health Ethics Committee in India (14/2020). Patients who meet the eligibility criteria will be approached to obtain informed consent for enrollment.

## 6.3 Consent for publication

Not applicable.

## 6.4 Availability of data and materials

The data that support the findings of this study are available on request from author MJar. The data are not publicly available due to them containing information that could compromise research participant privacy/consent.

## 6.5 Competing interests

JM, CH, SK, AW, AB, LB, NB, MK, VR, MJoh, EL, AR, AM, TS, MJon have no conflicts of interest to disclose.

CP serves on advisory boards for AstraZeneca, Boehringer Ingelheim, Merck Sharp and Dohme and Novartis.

CJ serves on advisory boards for AstraZeneca, Boehringer Ingelheim, Chiesi, GlaxoSmithKline, Novartis and Sanofi-Genzyme.

VJ has received grants from Baxter Healthcare, Biocon and GlaxoSmithKline, and speaker fees/advisory board from AstraZeneca, Baxter Healthcare, NephroPlus, Sanofi - all outside the submitted work. All fees paid to the organization.

MJar is responsible for research projects that have received unrestricted funding from Amgen, Baxter, Bayer, CSL Behring, Eli Lilly, Gambro, and Merck Sharp and Dohme; has served on advisory boards sponsored by Akebia, AstraZeneca, Baxter, Bayer, Boehringer Ingelheim, Merck Sharp and Dohme and Vifor; serves on the Steering Committee for trials sponsored by Chinook, CSL Behring and Janssen; serves on a Steering Committee for an investigator initiated trial in COVID-19 disease with funding support from Dimerix; spoken at scientific meetings sponsored by Amgen, Janssen, Roche and Vifor; with any consultancy, honoraria or travel support paid to the institution.

## 6.6 Funding

CLARITY is supported by the Australian Government’s Medical Research Future Fund Respiratory Medicine Clinical Trials Research on COVID-19 2020 grant scheme (ref. no. APP2002277).

## 6.7 Authors’ contributions

MJa made substantial contributions to the conception and design of the CLARITY study. JM and MJon made substantial contributions to the formulation of the trial stopping rules and trial analyses. CH, SK, AW, AB, CP, LB, NB, TS, VJ and MJa drafted the protocol. All authors read, edited and approved the manuscript for submission.

## 6.8 Acknowledgements

We thank all the participants in the CLARITY trial and the doctors, nurses, pharmacists and research staff at participating sites who ensure the successful implementation of the trial. We thank the CLARITY Consumer Engagement Committee for their helpful input into trial design and participant-facing materials. MJa is supported by a Medical Research Future Fund Next Generation Clinical Researchers Program Career Development Fellowship. TS is supported by a NHMRC Early Leadership Award (MRFF119513). MJon is supported by a grant from Snow Medical Foundation.

We would like to acknowledge the following CLARITY team members;

<u>Steering Committee Executive:</u> Meg Jardine (Chair), Vivekanand Jha (Co-Chair), Abhinav Bassi, Louise Burrell, Carinna Hockham, Christine Jenkins, Sradha Kotwal, Carol Pollock, Angus Ritchie, Arlen Wilcox

<u>Steering Committee Members:</u> Ashfak Bangi, Ashish Bhalla, Jenny Heng-Chen Chen, Sanjay D’Cruz, Michael Dymock, Simon Finfer, Greg Fox, Mayur Garg, Harry Gibbs, Lalit Gupta, Santosh Kumar Nag, Mark Jones, Benjamin Kwan, Angela Makris, George Mangos, Jennifer Martin, James McGree, Andrew McLachlan, Matthew O’Sullivan, Eugenia Pedagogos, Jeffrey Post, Vinay Rathore, Thomas Snelling, Louisa Sukkar, Richard Sullivan, Gian Luca Di Tanna, Jason Trubiano, Sophia Zoungas

<u>Study Clinician:</u> Sradha Kotwal

<u>Scientific Leads:</u> Abhinav Bassi, Carinna Hockham, Sradha Kotwal

<u>Project Manager:</u> Arlen Wilcox

<u>Central Management Team:</u> Grace Balicki, Kirston Barton, Nikita Bathla, Alison Coenen, Michelle Cummins, Sedricx Fontanilla, Enmoore Lin, Martyn Ralph, Nuria Zamora

<u>Statistical Team:</u> James McGree, Mark Jones, Tom Snelling

<u>Data Safety Monitoring Board:</u> Katherine Tuttle (Chair), Jonathan Craig, Stephane Heritier, Allison Lambert

<u>Consumer and Community Engagement Committee:</u> David Morgan and others

## Appendix Time-to-event outcome

To explore whether an alternative outcome might yield a reduced sample size, a simulation study was conducted for the primary outcome of time-to-ventilation/death. Prior information on this outcome was sourced from the Recovery trial (ClinicalTrials.gov, NCT04381936) which yielded an empirical hazard function as shown in Figure 4. As can be seen, there is essentially a fairly constant decline over a 28 day period. However, there are some deviations from this trend. For example, there is a second ‘hump’ in week 2 possibly associated with cytokine storm in a subset of patients (may be 30% of those hospitalised). This might explain this small hump however, it may also be artefactual. In addition, in regards to the first ‘hump’, mechanistically, it is plausible that in order to be enrolled in the trial there could be a small ‘healthy volunteer’ type selection bias whereby anyone whose death is imminent is not randomised (hence the slight increase in hazard over the first few days). Overall, it was thought that these deviations were not large enough to materially affect the simulations when compared to a monotonically declining hazard, so this is what was considered in the simulation study.

**Figure 4:**
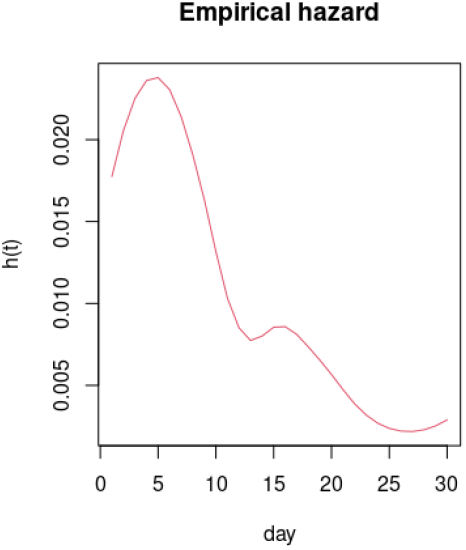
Empirical hazard function for patients with COVID-19 based on data from the Recovery trial (ClinicalTrials.gov, NCT04381936).

The empirical hazard function derived based on the results shown in Figure 4 is shown in Figure 5a. For comparison, a hazard function under the exponential distribution (i.e. constant hazard) and a Weibull distribution (i.e. monotonically decreasing) were also considered. These are shown in Figure 5b and c. If proportional hazards are assumed (between the placebo plus standard care and treatment plus standard care groups) then the hazard for the invention group can be defined via a proportionality parameter (*ψ*). This proportionality parameter reflects the treatment effect i.e. log *ψ* = *β*. All three hazard functions for treatment plus standard care group are also shown in Figure 5 with *ψ* = 0.87.

**Figure 5:**
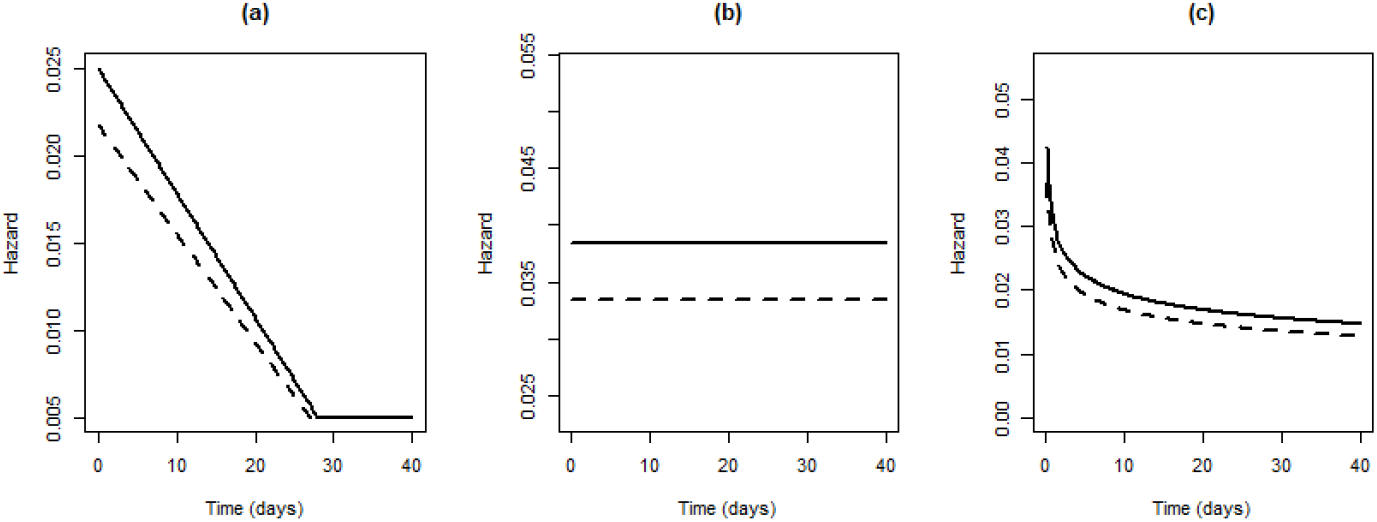
Proposed hazard functions for simulation study.

Once the hazard function is defined, then all functions needed to simulate time-to-event data (e.g. cumulative hazard, Survivor function, etc) can be derived. The approach for estimating power given a particular sample size is outlined in Algorithm 3, and is a standard simulation estimation approach. As can be seen, the hazard function (which may involve additional parameters *ϕ*), *ψ* and the sample size *N* are initially defined. Then, for a large number of iterations, treatment allocations are simulated and data are then simulated based on a probability density function and these allocations (line 3). A survival model is then re-fit to the data (line 4). Here, two types of survival models are fitted; a semi-parametric version where the baseline hazard is left undefined, and a Weibull proportional hazards model. The appeal of the semi-parametric model is that the baseline hazard is quite flexible, and should be able to capture the empirically defined hazard. The appeal of the Weibull model is that assuming a parametric hazard should yield gains in power, however the hazard may not be flexible enough to capture the assumed form of the empirical hazard. Both models were fit using maximum likelihood techniques. Based on the fitted model, a 95% confidence interval for log *ϕ* was found, and an indicator for whether this interval includes 0 was evaluated. Once this process was repeated a large number of times, the power was approximated by evaluating the proportion of times the 95% confidence interval for log *ϕ* did not include 0.

### Algorithm 3 Approach for estimating power for a given sample size with time-to-event outcome

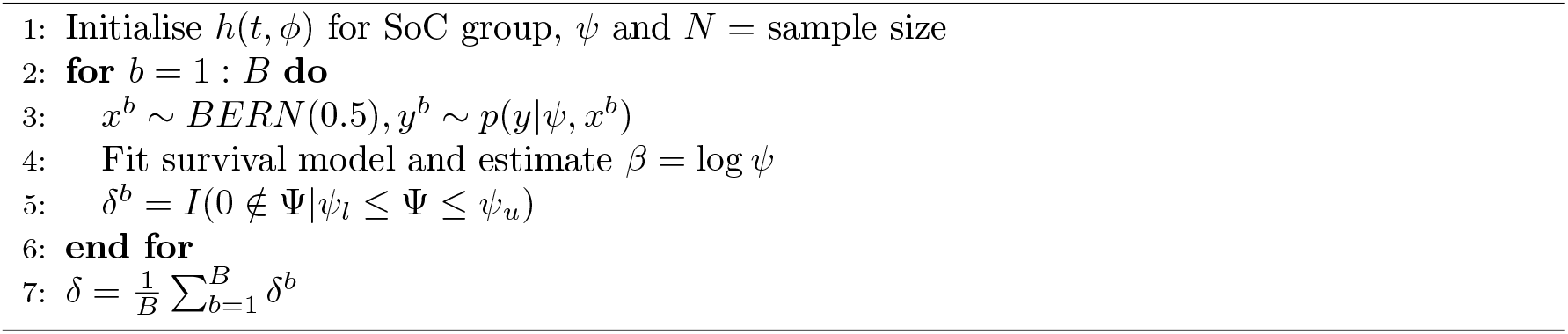

Approximate power for given sample sizes are shown in Figure 6 for the three hazard functions, *ϕ* = 0.87 and where the simulated data were refit under the semi-parametric and Weibull regression models. As can be seen, there is little difference in power estimates between the semi-parametric and Weibull models. However, when data were generated based on the empirical hazard, the maximum likelihood estimates for the Weibull regression model often failed to converge. Accordingly, only results for the semi-parametric model are shown for this case. Across all cases, it can be seen that a large number of participants need to be enrolled to achieve at least 60% or 80% power (particularly in the empirical and Weibull hazard function case). Given this, it was concluded that it was not worth pursuing a full Bayesian adaptive sample size implementation to explore required sample sizes across a variety of different settings.

**Figure 6:**
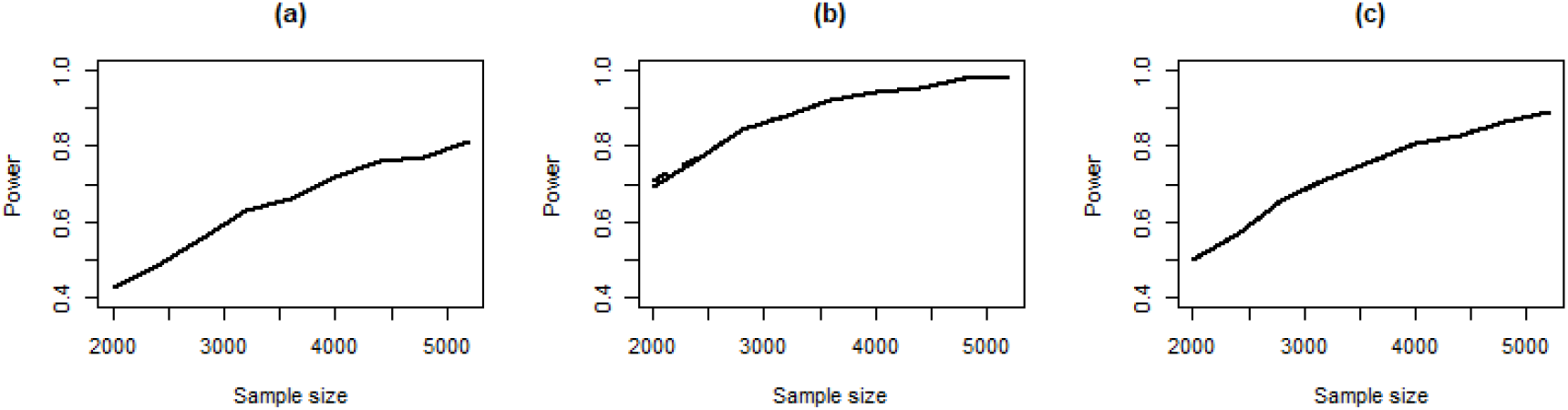
Approximate power for given sample sizes for *ϕ* = 0.87 and the (a) Empirical hazard function, (b) Exponential hazard function *λ* = 1/26, and (c) Weibull hazard function *γ* = 0.8, *λ* = 1/26 when the semi-parametric (–) and Weibull (– –) regression models were refit to the simulated data.

## Notes

### Clinical Trial

NCT04394117

### Author Declarations

Sydney Local Health District Ethics Review Committee

